## Supplementary File 1 for "Evaluating Large Language Models for Drafting Emergency Department Discharge Summaries"

**Background**

The November 2022 introduction of ChatGPT (GPT-3.5) and, subsequently, the more advanced GPT-4 has led to renewed focus on the use of natural language processing across a variety of domains.^1,2^ Large language models (LLMs) possess a range of capabilities which may be applied to the clinical domain, one of which is text summarisation. Writing concise but accurate summaries of clinical information is an important task within healthcare, allowing pertinent information to be communicated between medical professionals and from clinician to patient.

Patient discharge summaries, created following both hospital admission and Emergency Department visits, are an essential part of patient care, serving as a critical method of patient information transfer and providing instructions for the ongoing management of their illness.^3–5^ However, this task is time consuming and, often, discharge summaries are not completed in a timely manner.^5,6^ This is problematic given that the timeliness of discharge summary availability has been found to be associated with patients’ readmission rates, with the absence of a discharge summary associated with a 79% increased rate of 7 day readmission and 37% increased rate of 28 day readmission.^6^ In addition, given the increasing burden on healthcare providers using electronic health record (EHR) systems, there is a need to explore technological solutions to reduce physician burnout.^7^

Here, we explore the use of the GPT (GPT-3.5 and GPT-4) models to create Emergency Department discharge summaries. We evaluate model performance across three critically important areas: accuracy of summarisation, omission of clinically relevant information and presence of hallucinations in summarised text.

**Methods**

In this study, we seek to evaluate the ability of two widely used LLMs, GPT-3.5 and GPT-4, to summarise clinical information for patients discharged from the Emergency Department (ED). An initial dataset will be created based on the below inclusion criteria.

***Inclusion/exclusion criteria:***

- Adult (≥ 18 years) patients attending the ED with an available Emergency Medicine (EM) physician note
- Only patients who were subsequently discharged from the Emergency Department (rather than admitted to hospital) will be included
- The first EM physician note from each patient’s ED encounter will be retrieved based on note chart time. In cases where more than one note has been submitted at the same chart time, the longest note will be selected.
  - Only ED visits with notes containing all of 1) Chief complaint, 2) Physical Exam and 3) Initial assessment/ED course will be included
  - ED visits with notes >3500 tokens in length will be excluded due to the 4096 token limit of the GPT-3.5-turbo model

Two separate n = 100 random samples will be selected from this initial dataset. The first (aka the *development* set) will be used for prompt engineering attempts and for manual reviewer training, while the second sample (aka the *test* set) will be used for evaluation of GPT model summarisation performance.

The notes in this sample will be manually checked to confirm the note represents a valid History and Physical examination (H&P) note, detailing the patient’s ED course (including Presenting complaint, History of Presenting Complaint, Physical Examination findings, Investigation(s) performed and Impression/Plan).

Using the secure, HIPAA-compliant, UCSF Versa Application Programming Interface (API), we will prompt both GPT-3.5 and GPT-4 to summarise the provided ED physician note into a DC summary. The following prompt will be used, followed by the corresponding note for each patient, denoted by triple quotation marks:

- *“You are an Emergency Department physician. Below is the History and Physical Examination note for a patient presenting to the Emergency Department who was subsequently discharged. Write a discharge summary for the patient based on this note. Do not include any additional information not present in the note. \n\n """* Note text *""" ”*

The GPT-3.5- and GPT-4-generated completions will be collated and provided to two independent reviewers for manual evaluation. Reviewers will consist of a team of 5 Emergency Medicine (EM) residents, with an attending EM physician acting as the 3^rd^ party adjudicator for instances of disagreement among resident reviewers.

***Workflow***

Reviewers will be provided this protocol, including the detailed instructions (Appendix A, below) for review. An initial meeting will be scheduled following distribution of the protocol to provide an opportunity for reviewer queries about the protocol, associated instructions, or anything else to be addressed. Reviewers will then review an initial 15 samples prior to a second meeting, during which any additional queries (which arise following review of the first 15 samples) can be addressed and this protocol/instructions for reviewers modified accordingly. Subsequently, the remainder of the sample will be evaluated as per the protocol/instructions for reviewers. In the event that the protocol/instructions for reviewers require significant modification following initial review of the first 15 samples, these samples will be re-evaluated according to the modified protocol/instructions for reviewers.

***Timeline***

The proposed timeline for this project is as follows:

- Day 0: Distribution of protocol
- Day 2-4: Initial meeting
- Day 4-7: Review of first 15 cases
- Day 7: Second meeting; any subsequent queries addressed
- Day 7 – 21: Review of remaining cases +/- re-annotation of initial 15 cases based on feedback from Second meeting
- Day 21: Return of completed annotations
- Day 21 – 28: Analysis of results
- Day 28 – 35: Write up of results and submission to peer-reviewed journal

***Evaluation criteria***

The following three criteria will be evaluated.

- Criteria 1: Accuracy of GPT-summarised information in the DC summary
- Criteria 2: Hallucination of information in the summarised DC summary by GPT; and
- Criteria 3: Omission of relevant clinical information from the DC summary.

In order to perform a comprehensive evaluation, each of the below sections pertaining to a patient’s clinical history, examination and plan will be evaluated:

- Presenting complaint
- History of presenting complaint
- Past medical history
- Allergies/contraindications
- Review of systems
- Examination findings
- Blood test results
- Radiological investigations
- Plan
- Other notable events during ED stay (outside of above fields)

***Analysis***

After completion of DC summary evaluation by resident reviewers, responses will be collated and discordance between the two reviewers will be evaluated. Any disagreements will be evaluated by the 3rd party attending EM physician.

Evaluation results will be summarised in a descriptive analysis, detailing the percentage of DC summaries with inaccurate information, hallucinations and/or omission of relevant clinical information across the different subsections of the H&P note. Comparison of performance differences between GPT-3.5 and GPT-4 will be made, along with the overall word count of the GPT-generated discharge summaries.

Depending on the time taken to evaluate the first initial n = 100 sample, we may consider creating a second sample for manual review to increase the total sample size. We recognise that, for these types of study, larger samples may be more representative, but this must be balanced with the significant person-hours required for manual evaluation of the sample. Hence, we will make a decision on whether to increase the sample size once the size of the effort required to complete evaluation of the first n = 100 sample is known.

**Appendix A**

**Instructions for reviewers**

Please review the above protocol to familiarise yourself with the study background, aims, methods and timeline.

We will provide a simple interface on RedCap with which 1) the original ED note, 2) the GPT-3.5-generated DC summary and 3) the GPT-4-generated DC summary can be displayed, alongside a checkbox system for evaluating each of the below criteria.

*RedCap Guide (for evaluation/labelling)*

1. Login to redcap.ucsf.edu (using the username/password sent to your UCSF email address) and navigate to the ‘GPT_ED_summarisation_incl_notes_PROD’ project.
2. I have setup RedCap in the following way:
   a. Each resident reviewer should create *only 1 record*. After creating a new record, you will see a table with ‘NEW Record ID ##’ as the header – make a note of which number record you are (and **only fill in entries for this record number**)
   b. Within each record, all 100 cases are included – these are labelled *Index 0 to 99*
   c. I will give each reviewer 40 cases to review – the case numbers I give you should match the *Index ##.*
   d. For each case number you have been provided, click on that Index ## in the record (e.g click on the white circle under ‘Status’ to bring up the case). This will bring you to a screen that contains the ‘Original ED provider note’ along with GPT-3.5 and GPT-4 generated summaries. Fill in the corresponding *Index ##* for each case (this is just a sanity check to make sure the case you think you are doing matches the case in the record).
      i. E.g if given cases 0-9, fill in *Index 0, Index 1, .. Index 9*
   e. Field-by-field instructions for completing each form:
      i. Index ID: *write the case # here to allow identification of the case following export of all data when annotation has been completed*
      ii. Annotator initials: *write your initials*
      iii. Index (hardcoded): *this is hardcoded (can’t be edited) and should match the Index ID you have input, above*
      iv. Encounterkey (hardcoded): *please* *ignore this field – it contains the encounter ID for the ED note and is for my reference*
      v. Original ED provider note: *Contains the input text for the GPT model, including both a) the GPT prompt and b) the original note text*
      vi. GPT-3.5-turbo (chatGPT) generated DC summary: *Contains the chatGPT generated discharge summary*
      vii. GPT-4 generated DC summary: *Contains the GPT-4 generated discharge summary*
      viii. See the instructions below for completion of the True/False boxes, labelling first the GPT-3.5-turbo (ChatGPT) summaries, followed by the GPT-4 summaries
            1. When marking a response as TRUE, this should unlock a text box underneath, which will prompt you for ‘Reasons for marking response TRUE (short, single sentence, description)’
      ix. Form status
          1. *After completing all the required fuilds, Mark Form Status as *Complete* after completion* (this will make it green) on the dashboard
3. For those who prefer to dual screen (with note text on one screen and RedCap on the other screen), I will also send out the raw .txt files (in compressed .zip format; you just need to unzip them beforehand – make sure you only store these files on a UCSF device if downloading them locally), organised in folders where the folder ## corresponds to the case ##.
4. I looked into making the formatting of the original ED note more readable – unfortunately, the note has been flattened (i.e useful markers of new line and other formatting notation has been removed), so it isn’t possible to return it to a more readable format.

*Reviewer case ## assignment*:

- Alexa Lucas: 0 – 39 (please do 0-14 as the initial 15)
- Kishan Patel: 0 - 29, 40 - 49 (please do 0-14 as the initial 15)
- Terri Tang: 50 - 89 (please do 50-64 as the initial 15)
- Fiona Chen: 50 - 79, 90 - 99 (please do 50-64 as the initial 15)
- Karan Bains: 30 - 49, 80 - 99

*Evaluation of DC summaries*

Please answer the following questions to evaluate the quality of information present in the GPT-3.5/GPT-4 generated DC summaries provided. Use only the information available in the original ED note to inform your responses. For each note section (i.e Presenting complaint, History of presenting complaint etc), answer the following questions, marking the provided checkbox with an X for each TRUE response. Please also provide a short (single sentence) description of the reasons for marking a response TRUE.

Criteria 1: Inaccurate information

- *(For each of the below subsections) Does the DC summary contain* ***inaccurate*** ***information*** *(Y/N):*
  - Presenting complaint
  - History of presenting complaint
  - Past medical history
  - Allergies/contraindications
  - Review of systems
  - Examination findings
  - Blood test results
  - Radiological investigations
  - Impression/Plan
  - Other notable events during ED stay (outside of above fields)

Criteria 2: Hallucinations

- LLM hallucination refers to the phenomenon whereby an LLM generates seemingly realistic information that does not correspond to any real-world input – in other words, they are ‘made up’ outputs which sound plausible but are either factually incorrect or unrelated to the given context.
- Here, we distinguish hallucinations from inaccuracies (Criteria 1) by specifying that, for a GPT-generated information to be classified as hallucination rather than inaccurate information, its correct value must **not** be present in the original text.
  - For instance, if the original note states ‘Reviewed a 52 yo M patient with chest pain’ and the GPT model summarises this as ‘Reviewed a *42* yo male patient with chest pain’, this would be classed as *inaccurate* information.
  - In contrast, if the original note states ‘Reviewed a 52 yo patient with chest pain’ without mention of the patient’s sex (anywhere in the note), and the GPT model summary states ‘Reviewed a 52 yo *male* patient with chest pain’, this would be deemed a hallucination as the patient’s sex has been ‘made up’ aka hallucinated by the GPT model.
- *(For each of the below subsections) Does the DC summary contain* ***hallucinations*** *(Y/N):*
  - Presenting complaint
  - History of presenting complaint
  - Past medical history
  - Allergies/contraindications
  - Review of systems
  - Examination findings
  - Blood test results
  - Radiological investigations
  - Impression/Plan
  - Other notable events during ED stay (outside of above fields)

Criteria 3: Omission of clinically relevant information

- We recognise that there may be differences of opinion as to what information within a patient’s symptoms, past medical history, examination findings, investigation results and follow up plan should be included in a DC summary. In contrast to the above two criteria, we consequently expect there to be greater discordance between the two reviewers for Criteria 3.
- We ask reviewers to evaluate Criteria 3 based on their own clinical practice. We propose that cases where information has been omitted by GPT-3.5 and/or GPT-4 that the reviewer deems relevant (either for the patient’s information, for the information of future healthcare providers, or for any other reasons) to be included in the DC summary should be recorded with an ‘X’ (TRUE) in the checkbox and a short description should be provided.
- Discordance among reviewers, as with the above two criteria, will be resolved by the 3rd party EM attending reviewer.
- *(For each of the below subsections) Does the DC summary omit clinically relevant information (Y/N):*
  - Presenting complaint
  - History of presenting complaint
  - Past medical history
  - Allergies/contraindications
  - Review of systems
  - Examination findings
  - Blood test results
  - Radiological investigations
  - Impression/Plan
  - Other notable events during ED stay (outside of above fields)

**Examples (for each of the above criteria):**

- Criteria 1 [inaccurate information] example: (See example_note 0): If a patient’s note says ‘Allergies: Penicillin’, and GPT-3.5 summary has written ‘NKDA’, this would be classified as inaccurate information (Criteria 1), and the checkbox next to ‘Allergies/contraindications’ should be marked with an ‘X’ (TRUE), with ‘States “NKDA” but patient has penicillin allergy’ provided as the explanation.
- Criteria 2 [hallucination] example: (See example_note 2): ‘He was advised to avoid any allergens, including aloe and penicillins.’ – there was no such advice in the text, where the only mention of Aloe is to document that the patient has an aloe allergy.
- Criteria 3 [omission of clinically relevant information] example: (See example_note 3): Patient presented with trauma (bicycle vs car), went over handlebars and hit head but ‘without loc and states he remembers event’. GPT-4’s DC summary reported that the patient did not lose consciousness, but omitted to mention that he ‘stated he remembers event’. In my practice, I usually like to document ‘patient remembers events leading to..’ as objective evidence that they did not lose consciousness, and therefore I would mark this as TRUE for the ‘History of Presenting complaint’ section, with comment – ‘No mention of patient remembering event as confirmation of no LoC’.
