## Supplementary File 2 for "Evaluating Large Language Models for Drafting Emergency Department Discharge Summaries"

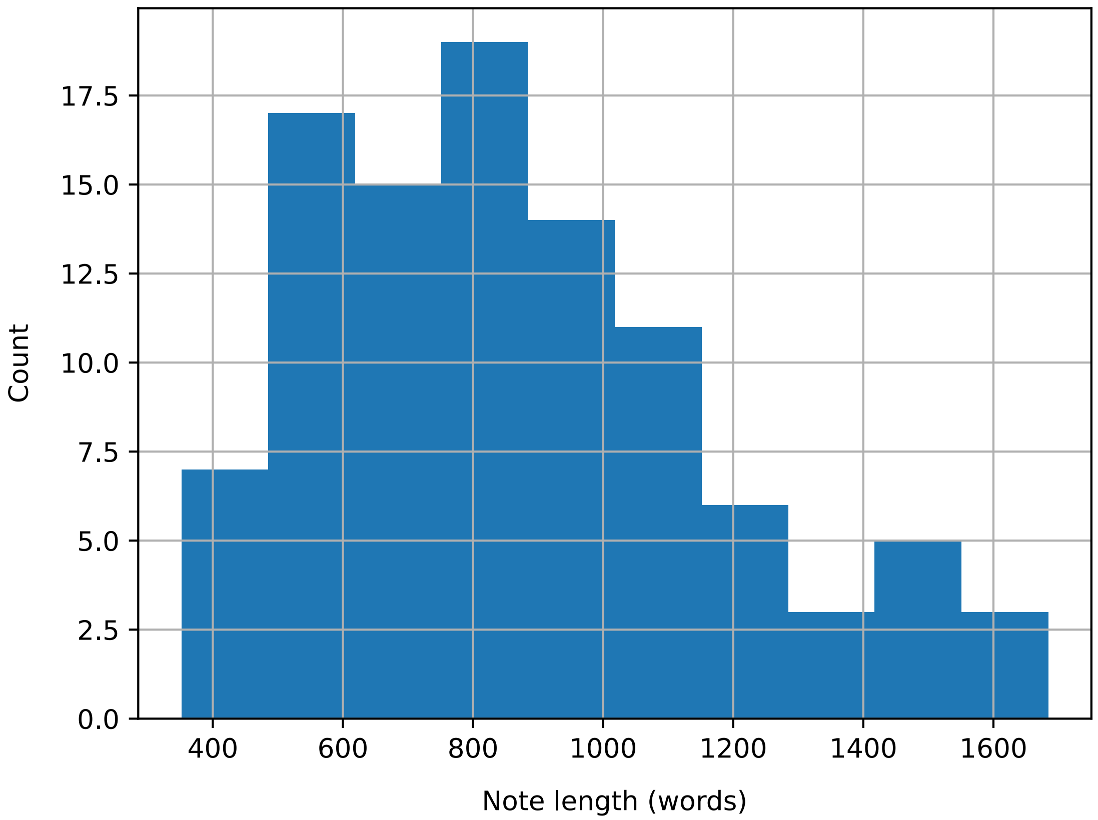


**Figure S1.** Histogram of original Emergency Medicine provider note length among the n = 100 sample of Emergency Department encounters randomly selected for GPT-3.5-turbo and GPT-4 summarization.

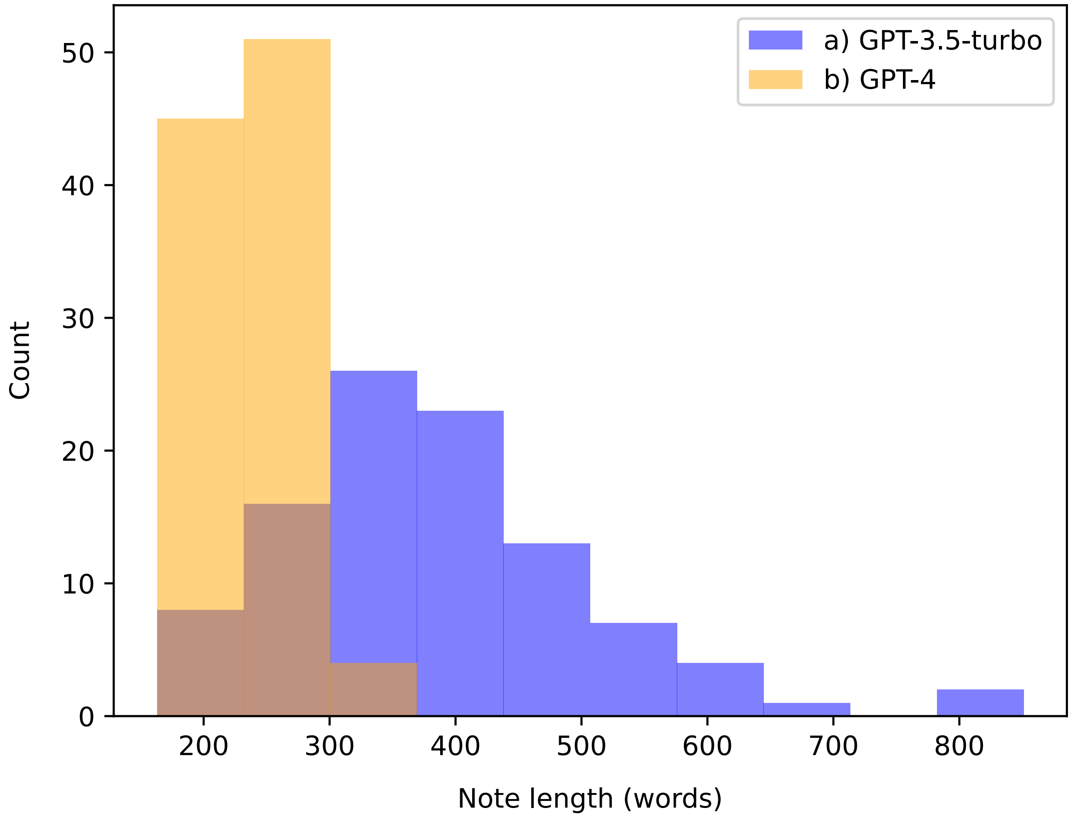


**Figure S2.** Histogram of word counts of a) GPT-3.5-turbo and b) GPT-4 generated discharge summaries.

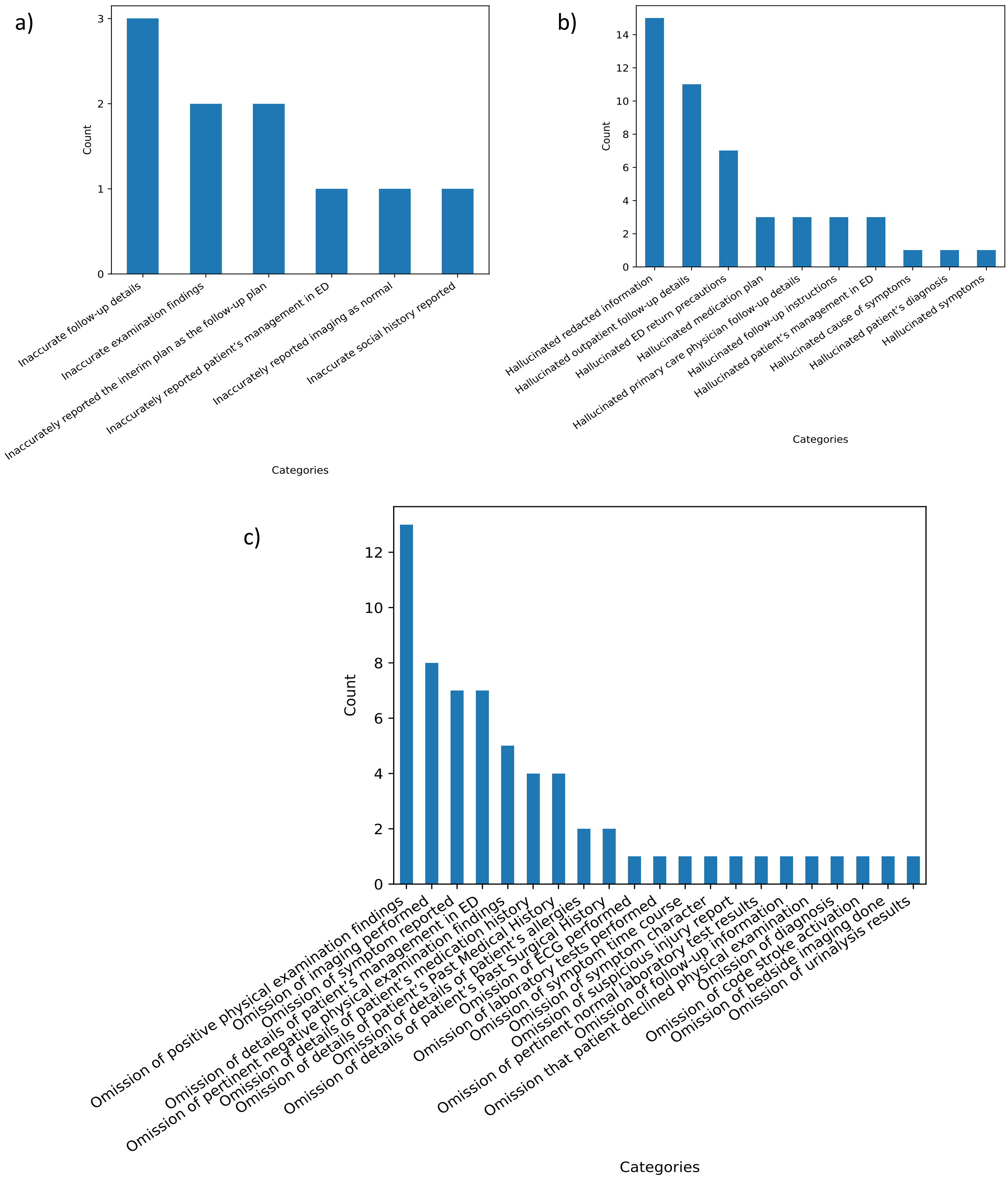


**Figure S3.** Manual categorization of reviewer comments providing further details for each error subtype [a) Inaccuracy, b) Hallucination, and c) Clinical omission] among GPT-4-generated discharge summaries compared to the ground-truth, original Emergency Medicine provider note.

a)

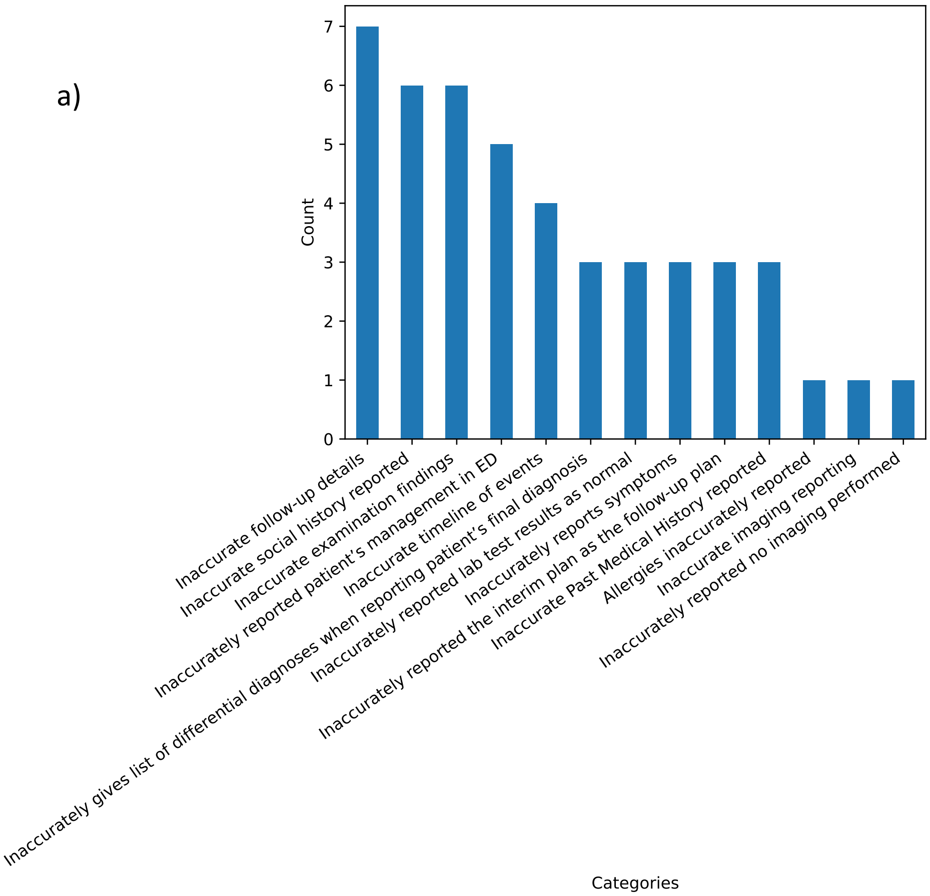

Count
Inaccurate follow-up details
Inaccurate social history reported
Inaccurate examination findings
Inaccurately reported patient's management in ED
Inaccurate timeline of events
Inaccurately gives list of differential diagnoses when reporting patient's final diagnosis
Inaccurately reported lab test results as normal
Inaccurately reports symptoms
Inaccurately reported the interim plan as the follow-up plan
Inaccurate Past Medical History reported
Allergies inaccurately reported
Inaccurate imaging reporting
Inaccurately reported no imaging performed
Categories


b)

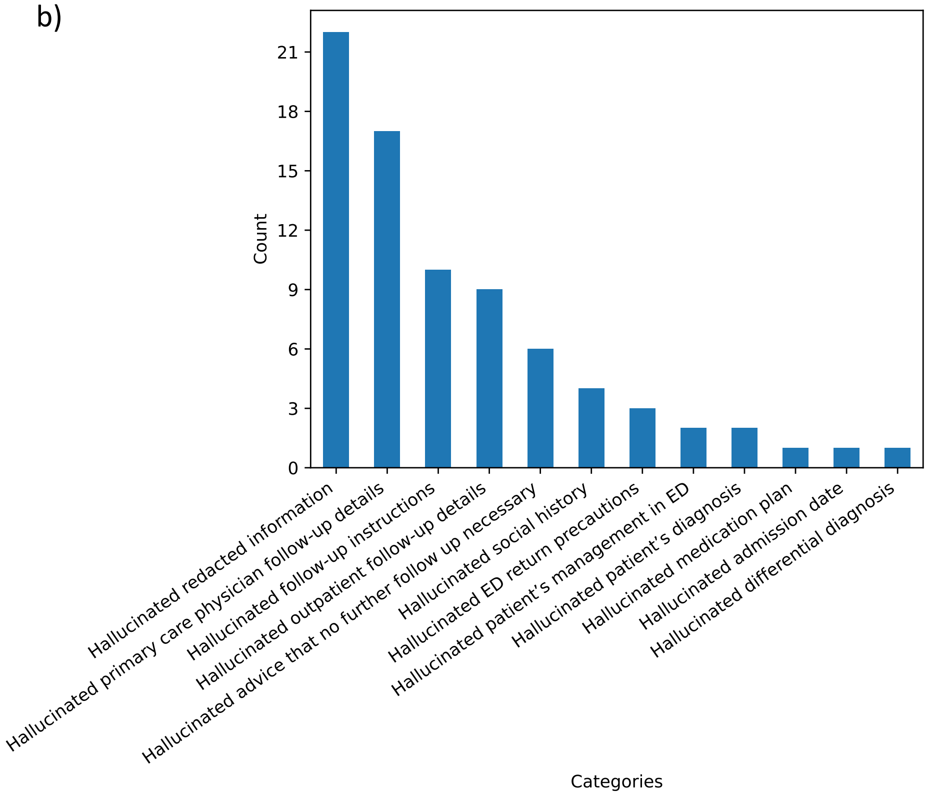

Count
Hallucinated redacted information
Hallucinated primary care physician follow-up details
Hallucinated follow-up instructions
Hallucinated outpatient follow-up details
Hallucinated advice that no further follow up necessary
Hallucinated social history
Hallucinated ED return precautions
Hallucinated patient's management in ED
Hallucinated patient's diagnosis
Hallucinated medication plan
Hallucinated admission date
Hallucinated differential diagnosis
Categories

c)

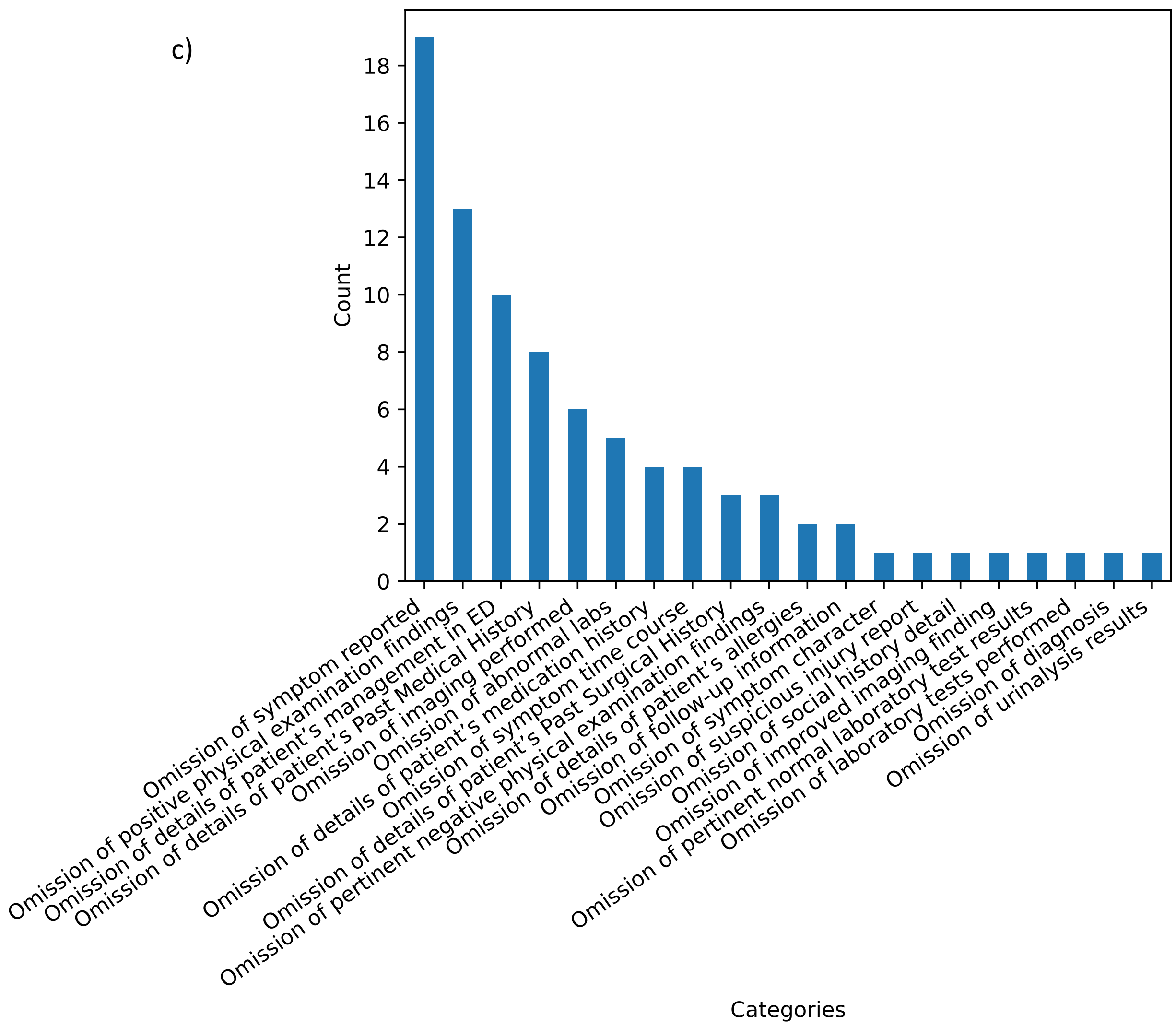


**Figure S4.** Manual categorization of reviewer comments providing further details for each error subtype [a) Inaccuracy, b) Hallucination, and c) Clinical omission] among GPT-3.5-turbo-generated discharge summaries compared to the ground-truth, original Emergency Medicine provider note.

| | Number of labels with inter-reviewer agreement (%) | | |
|---|---|---|---|
| | Inaccuracy | Hallucination | Omission |
| GPT-3.5-turbo summaries | 936/1000 (93.6%) | 917/1000 (91.7%) | 906/1000 (90.6%) |
| GPT-4 summaries | 979/1000 (97.9%) | 954/1000 (95.4%) | 931/1000 (93.1%) |
| **Total*** | **1915/2000 (95.8%)** | **1871/2000 (93.6%)** | **1837/2000 (91.9%)** |

**Table S1.** Initial inter-reviewer agreement rates by error type, prior to consensus agreement. *$p < 0.001$ (Chi-squared test, $\chi^2 = 26.0$).

| Error Type | Error category | Example reviewer comment* | Count |
| --- | --- | --- | --- |
| Inaccuracy | Inaccurate follow-up details | "There was no plan [in the original note] for *her* to set up an appointment with her personal neurologist; [the original note's] plan is for a neurology referral" | 7 |
| | Inaccurate social history reported | "Patient does use alcohol, just rarely [GPT summary incorrect states the patient doesn't use alcohol]" | 6 |
| | Inaccurate examination findings | "[GPT summary] contradicts itself: [in the GPT summary, there is an] abnormal bimanual exam reported in ED course (which would otherwise be okay), but the Physical Examination section reports that the systems (including the genitourinary system) were all 'within normal limits' – hence inaccurate" | 6 |
| | Inaccurately reported patient's management in ED | "[GPT summary] says patient was discharged with rx of tylenol; rather patient was offered dose in the ED but refused" | 5 |
| | Inaccurate timeline of events | "[Original note] says he had similar symptoms 6 months ago, but [GPT] summary says several years ago" | 4 |
| | Inaccurately gives list of differential diagnoses when reporting patient's final diagnosis | "GPT summary reports 'the differential diagnosis included ascites due to decompensated cirrhosis, esophageal variceal bleeding, and HCC with tumor thrombus in the portal vein' but the patient came in with worsening abdo distention due to ascites (so variceal bleed wasn't an issue)"; "[GPT summary] reports differential as diagnosis – 'patient was diagnosed with strep pharyngitis, viral illness, bronchitis and possible pneumonia'" | 3 |
| | Inaccurately reported lab test results as normal | "[GPT summary] stated Hgb and Hct were within reference range though were low" | 3 |
| | Inaccurately reports symptoms | "GPT writes 'epistaxis followed by rhinorrhea/post-nasal drip', however [original] note only states he's currently experiencing rhinorrhea/drip not that it was sequentially followed by the epistaxis" | 3 |
| | Inaccurately reported the interim plan as the follow-up plan | "[GPT summary] states [patient is] to be discharged with f/u plan for psych and SW, but this was part of the sign out to be done in the ED" | 3 |
| | Inaccurate Past Medical History reported | "GPT inaccurately says no PMH found, but note said he had HTN" | 3 |

| | | | |
|---|---|---|---|
| | Allergies inaccurately reported | "[Original note states] patient has shortness of breath to latex, not penicillin [as incorrectly stated in GPT summary]" | 1 |
| | Inaccurate imaging reporting | "[GPT] note inaccurately mentions 'the patient underwent a thoracentesis, which showed a decrease in the size of the right pleural effusion' – but it was the CXR which showed this following thoracentesis" | 1 |
| | Inaccurately reported no imaging performed | "[GPT summary] states 'no specific lab, imaging, or ECG results were mentioned in the note' [when in fact] CXR and KUB [were] ordered and results discussed in [the original] note | 1 |
| Hallucination | Hallucinated redacted information | "redacted portion after 'mechanical …' was filled in as 'injury' by GPT"; "hallucination that patient will be sent to treatment center, word "center" mentioned between multiple asterix'ed out portions"; "hallucinated redacted portions of losing job" | 22 |
| | Hallucinated primary care physician follow-up details | "Hallucinated PCP follow-up which was not originally listed"; "Hallucinated plan for symptom control and PCP follow-up (no mention in original note)" | 17 |
| | Hallucinated follow-up instructions | "Created follow-up instructions that were not previously mentioned"; "Entire follow up section is not present in original note" | 10 |
| | Hallucinated outpatient follow-up details | "Hallucinated follow-up plan with cardiology"; "Original note did not mention ortho follow up"; "No mention of further outpatient workup in [original] note" | 9 |
| | Hallucinated advice that no further follow up necessary | "[GPT] summary says "no other outpatient work up necessary" but this was not mentioned in the note, just said to follow up with PCP"; "'No further outpatient workup was deemed necessary' - never stated in original note"; "Final portion of note says no further outpatient w/u deemed necessary, which was never stated" | 6 |
| | Hallucinated social history | "[GPT summary] states [patient] denies drugs and smokeless tobacco but it's simply not listed [in the original note] so inaccurate to say he denies"; "Patient did not report meth and cocaine use in [original note] HPI." | 4 |
| | Hallucinated ED return precautions | "No specific return precautions discussed in non-redacted portion of note, did not say "if symptoms worsen" specifically"; "States pt stable for ED discharge and provides discharge instructions, however this is not | 3 |

| | | | |
|---|---|---|---|
| | | present in note - possibly was in redacted portion” | |
| | Hallucinated patient’s management in ED | “Patient's daughter’s involvement not mentioned in [original] note” | 2 |
| | Hallucinated patient’s diagnosis | “Original note did not say [patient had] likely migraine headache [which is stated in the GPT summary]” | 2 |
| | Hallucinated medication plan | “Hallucinates ‘advised to continue home meds, return precautions, etc.’” | 1 |
| | Hallucinated admission date | “Admission date is hallucinated, patient could have come in the day before.” | 1 |
| | Hallucinated differential diagnosis | “Lyme disease listed in [GPT summary’s] differential, not in note.” | 1 |
| Clinical Omission | Omission of symptom reported | “Left out lack of flatulence x 10 days”; “left out that en route had episode of emesis and required Zofran”; “Omitted watery stools”; “Omission of positive ROS for abdominal pain.”; “Lists all the negative ROS but not positive for SOB” | 19 |
| | Omission of positive physical examination findings | “No mention of sutures in place”; “Omitted portion of PE discussing healing small surgical incisions”; “Crackles and desaturations and foot numbness omitted.”; “+Abnormal gait, and use of cane omitted.” | 13 |
| | Omission of details of patient’s management in ED | “Omitted that patient was given morphine”; “Omitted IVF and tylenol from plan”; “Omission that dialysis was performed”; “Omitted Fleet [enema] for constipation” | 10 |
| | Omission of details of patient’s Past Medical History | “Omitted baseline deficits from stroke”; “Omitted healed old neck lacerations (self-harm), relevant as pt presented intoxicated”; “[Omitted] history of CAD s/p CABG - relevant in patient presenting with chest pain” | 8 |
| | Omission of imaging performed | “Did not mention XR”; “Omitted CT scan performed”; “Omitted CXR” | 6 |
| | Omission of abnormal labs | “Omission of AKI”; “[Omitted] leukocytosis with small left shift”; “Omitted hyponatremia finding” | 5 |
| | Omission of details of patient’s medication history | “Omitted estrogen use and immunosuppression”; “Omitted antibiotics already given for UTI and persistent symptoms” | 4 |
| | Omission of symptom time course | “Omits timing of sudden onset 2 hours ago”; “Omitted fact that g-tube was dislodged 2 hours ago”; “Omitted timeline of symptoms, 3 hour epistaxis” | 4 |
| | Omission of details of patient’s Past Surgical History | “[Omitted] throat cyst removal surgery, history of ovarian cysts”; “Omits history of PEG”; “Omitted cholecystectomy surgery” | 3 |

| | | | |
|---|---|---|---|
| | Omission of pertinent negative physical examination findings | "Hand exam needs more detail including [the fact there is] no tendon or joint involvement, that the lac was explored deeply"; "No mention of pertinent negative of head impulse test results (which indicates peripheral pathology)"; "Would include lack of fluorescein uptake as important finding" | 3 |
| | Omission of details of patient's allergies | "Left out contraindication for no BP on right arm s/p mastectomy"; "Does not mention allergies" | 2 |
| | Omission of follow-up information | "Omitted c/f slow subdural development and need for repeat CT" | 2 |
| | Omission of symptom character | "Omitted description of abdo pain as intermittent epigastric, burning/sharp/crampy, non radiating, and association with stools" | 1 |
| | Omission of suspicious injury report | "Omitted suspicious injury report" | 1 |
| | Omission of social history detail | "Omitted she is wheelchair user" | 1 |
| | Omission of improved imaging finding | "Omitted decreased ascites" | 1 |
| | Omission of pertinent normal laboratory test results | "Left out normal APAP level" | 1 |
| | Omission of laboratory tests performed | "Omitted discussion of labs" | 1 |
| | Omission of diagnosis | "Did not include the presumptive diagnosis selection (menstrual cramps) amongst the various differential diagnosis entities" | 1 |
| | Omission of urinalysis results | "Omitted positive urine drug screen for cocaine (not extremely relevant)" | 1 |

**Table S2.** Manual categorization of reviewer comments providing further details for each error subtype among GPT-3.5-turbo-generated discharge summaries compared to the ground-truth, original Emergency Medicine provider note. *Comments reported with minor modifications to syntax for improved readability.
